# Multimodal predictors of disability progression and processing speed decline in relapsing-remitting multiple sclerosis

**DOI:** 10.1101/2025.02.09.25321961

**Authors:** Max Korbmacher, Ingrid Anne Lie, Kristin Wesnes, Eric Westman, Thomas Espeseth, Karsten Specht, Ole Andreas Andreassen, Lars Tjelta Westlye, Stig Wergeland, Kjell-Morten Myhr, Øivind Torkildsen, Einar August Høgestøl

## Abstract

The underlying mechanisms for neurodegeneration in multiple sclerosis are complex and incompletely understood. Multivariate and multimodal investigations integrating demographic, clinical, multi-omics, and neuroimaging data provide opportunities for nuanced analyses, aimed to define disease progression markers. We used data from a 12-year longitudinal multicenter cohort of 88 people with multiple sclerosis, to test the predictive value of multi-omics, T_1_-weighted MRI (lesion count and volume, lesion-filled brain-predicted age), clinical examinations, self-reports on quality of life, demographics, and general health-related variables for future functional and cognitive disability. Systematic increases in Expanded Disability Status Scale (EDSS) scores were used to stratify a progressive disability group (PDG) from relatively stabile disability. A processing speed decline group (PSDG) was defined by a ≥20% decrease from the maximum (cognitive) Paced Auditory Serial Addition Test score. We used a multiverse approach to identify which baseline variables were most predictive for PDG and PSDG memberships, considering multiple analysis paths.

Future disability (median area under the curve: mAUC=0.83±0.04, median Brier score: mBS=0.16±0.02) and the loss of processing speed (mAUC=0.89±0.05, mBS=0.10±0.03) could be successfully classified across models. Varibles significantly (median p-values<0.05) predicting stable disability included receiving disease modifying treatment at 12-year follow-up (median Odds Ratio: mOR_PDG_=7.44±4.07, p_median_=0.013, proportion of the OR’s directionality: PORSD=100%), lower baseline EDSS for each 1-unit (mOR_PDG_=0.25±0.11, p_median_=0.013, PORSD=100%), and counter-intuitively every year increase in baseline age (mOR_PDG_=1.12±0.04, p_median_=0.020, PORSD=100%), and lower vitamin A per 1 umol/L (mOR_PDG_=0.10±0.05, p_median_=0.016, PORSD=99.7%) and D levels per 1 nmol/L (mOR_PDG_=0.95±0.02, p_median_=0.025, PORSD=100%). Variables significantly predicting stable processing speed were receiving disease modifying treatment at 12-year follow-up (mOR_PSDG_=0.10±0.08, p_median_=0.013, PORSD=100%) and baseline PASAT score (mOR_PSDG_=0.86±0.03, p_median_=0.005, PORSD=99.73%). These findings were supported by an additional simulation study.

Concordant with the literature, disease modifying treatments influence disability progression, as well as a higher EDSS and PASAT scores at measurement start. Experimental and counterintuitive findings on vitamin A and D levels require further validation. The large variability across models suggests a strong influence of analytic flexibility, such as the selection of covariates.

## Introduction

Multiple sclerosis (MS) is a chronic immune-mediated disease of the central nervous system. Globally, there are more than 2.8 million people with MS (pwMS), with a higher prevalence in regions such as North America and Europe compared to Africa and East Asia.^1^ Despite significant advancements in disease-modifying therapies over the last two decades, pwMS exhibit considerable heterogeneity in their disease progression and response to treatment.^2,3^ While some pwMS develop severe disability despite high-efficacy disease-modifying therapies, others remain stable under similar regimens.^4^ This variability underscores the importance of stratifying pwMS to identify distinct disease trajectories and improve treatment precision.

Disease trajectories can be defined by disease progression or processing speed decline , which have been described to be predictive of each other,^5,6^ with processing speed potentially reflecting a unique dieasese dimension.^7,8^ A clinically meaningful definition of disease trajectories requires a multimodal approach that integrates clinical, imaging, and molecular markers.^9^

An advantage of a multivariate approach is the possibility of highlighting multiple variables associated with MS, both those assessed in routine clinical assessments and beyond.^10^ For example, various brain imaging markers such as lesion count^11^, lesion volume^12,13^, and brain age^14^ have been linked to disability. Brain age is an age estimate based on a comparison of a person’s brain MRI with a large dataset from a healthy population.^15^ Furthermore, several serum markers have been explored as prognostic markers in MS, including serum neurofilament light chain (NfL)^16^, chitinase-3-like protein1 (CHI3L1)^17,18^, vitamin D^19^, and potentially other vitamins^20,21^, and HLA-DRB1 carriership^19^. General health risk markers such as smoking^22^ and high body mass index (BMI)^23^ also contribute to MS-related outcomes. Clinical variables such as the relapse rate^24^, Expanded Disability Status Scale (EDSS)^25^, and cognitive performance, for example measured by the Paced Auditory Serial Addition Test (PASAT)^26^, further enhance the understanding of disease-related impairment. Additionally, patient-reported outcome measures (PROMs)^27^ can offer valuable insights for a more comprehensive assessment of disease impact.

We have established a unique cohort, containing longitudinal recordings for all the above variables. While the richness of such data provides insights from many different perspectives, it also presents analytical challenges, particularly the potential for bias introduced by researchers’ subjective decisions in data (pre)processing, modelling, and interpretation. Study designs often highlight one or few predictors of MS outcomes, not accounting for multiple other predictors and their possible combinations. However, due to statistical dependencies, the selection of the variables included in such models influences the observed results, in addition to many other decision researchers make when processing and analysing data.^28^

To enhance reproducibility and minimize bias, robust and systematic analytical frameworks are essential.^28^ Multiverse analyses, which explore multiple plausible analytical paths and evaluate their impact on results, offer a promising approach to address analysis bias.^29^

Relevant not only to MS research, variable selection remains an unsolved statistical issue^30^ for which multiverse analyses provide an opportunity to decrease analysis-specific bias when presenting which variables are important for disease trajectories. In our study, we combine commonly used data from clinical follow-up and experimental biomarkers of MS derived from MRI, blood samples, clinical assessments and quality of life (QoL). We stratify pwMS into groups of cognitive and functional impairment and showcase variables predicting future trajectories already at baseline.

## Materials and methods

### Sample

The sample included 88 pwMS with relapsing-remitting MS (RRMS) participating in the omega-3 fatty acid in MS (OFAMS) multicentre clinical trial^31^. The trial entailed data collection every 6 months over 2 years, followed by a single follow-up visit 10 years after the original trial concluded. A total of 85 (97%) of the original clinical trial participants were included and considered in the present analyses. In addition to age, sex, BMI, smoking status, and ω-3 supplementation (original clinical study intervention), the collected data used in this study entailed assessments of clinical (EDSS)^25^ and processing speed (PASAT)^26^, together with QoL, evaluated by the short form quality of life inventory (SF-36)^32^, T_1_- and T_2_-weighted MRI and blood samples (see Supplemental Table 1 for overview over variables; Supplemental Figure 1 for variability of raw measures). All participants were treated with inferon beta-1a starting at month 6 after inclusion in the study, but were treatment naïve before that. As new medications were introduced, the participants’ treatment plans changed unsystematically at various points with ultimately 62 of 85 of the included pwMS using disease modifying treatment.

We also used a reference MRI sample (see Supplemental Table 2) to establish brain age models (n_training_=58,317, n_validation_=6,608, age span = 5-90), non-clinical repeatedly sampled data of three individuals across one year (minimum of 25 scans each, Bergen Breakfast Scanning Club [BBSC])^33^, and a cross-sectional MS sample (n=748, mean age = 38.63±9.46 years) to validate the brain age model in a relevant clinical cohort, matched with healthy controls (n=751, mean age = 38.73±9.61).

### Demographic, clinical, self-reported, multi-omics, and neuroimaging assessments

Age, sex, BMI, and smoking status (smoker/non-smoker assessed by both serum cotinine levels and self-reports) were general variables available for all participants. Clinical and cognitive scores included EDSS^25^ and PASAT^26^, and the number of relapses 12 months prior to study start. SF-36 indicated QoL in eight categories [physical functioning, role limitations due to physical health, role limitations due to emotional problems, energy/fatigue, emotional well-being, social functioning, pain, and general health] based on the National Multiple Sclerosis Society’s scoring recommendations^34,35^ with scores ranging from 0-100 (worst to best outcome) except for general health (1-5). Serum analyses for the current sample^16,36^, and MRI acquisition and post-processing (cortical reconstruction and lesion segmentation) have been described previously^22^. MRI scanners and sequences can be found in Supplemental Table 3.

MRI markers of interest included baseline brain age gap (BAG) in years (see next section) from T_1_-weighted MRI data, as well as lesion count and volume (mm^3^) from T_2_-weighted MRI data. Serum markers included CHI3L1 (mg/ml), NfL (pg/ml), and vitamin A and E levels (in µmol/L) and vitamin D levels (in nmol/L), as well as HLA−DRB1 carriership (yes/no). Intra-class correlation coefficients were good (ICC > 0.82), apart from CHI3L1 (ICC ≈ 0), vitamin D levels (ICC = 0.56), and zero-variance variables (see Statistical Analyses). As a consequence of the instability of CHI3L1, we excluded the marker from the analyses.

### Sample stratification

#### EDSS-based stratification

We first stratified pwMS into a progressive disability group (PDG) and the rest of the participants into a stable or improving disability group (SIDG). As previously discussed,^37^ PDG (n=37, 42%) was defined by progressive disability consisting of at least 1.5-point EDSS increase, when baseline EDSS was 0; 1.0-point increase when baseline EDSS was between 1.0 and 5.5; and 0.5 increase when baseline EDSS was equal or more than 6.0, with a follow-up time of 128-156, M=141.93±5.83 months.

#### PASAT-based stratification

In a second, independent stratification, we defined a processing speed decline group (PSDG) based on a 12-point decrease in PASAT score (corresponding to 20% of the maximum score of 60 points, which was informed by previous literature^38^) either between baseline and 144-month follow-up or 24-month session and 144-month follow-up, and with the scores decreasing over time (n_PSDG_=22). In contrast, a stable processing speed group (SPSG) was defined by smaller processing speed decline than in the PSDG, within expected effects of aging, or even stable or improving PASAT scores.

#### Brain age

We developed a machine-learning based generalized additive model (allowing polynomials up to the 4th order) trained on 58,317 healthy controls’ cortical thickness, volume and surface area averages across Desikan-Killany atlas^39^ parcels to predict age. Such models allow sufficient flexibility to approximate non-linear age-dependencies, while still being simple enough to be interpretable.

Bias corrections for brain age prediction models are important as brain ages are otherwise systematically overestimated for lowest ages and underestimated for highest ages, respectively. While this is a know problem, previous studies do usually only apply post-hoc corrections, which do however not remove all of the bias. Hence, we used a combined approach: The original training sample (n=58,317) was up-sampled (to n=150,517) using SMOGN^40^ to approximate a unimodal distribution to minimize training sample age-distribution bias. To account for remaining age-distribution bias, we applied a post-hoc correction using the training sample’s linear age and brain age (BA) association’s intercept (a) and slopes (b): Corrected BA = BA + (Age - (a+b * BA)). The correction procedure improved model performance (Supplemental Table 4). Finally, the brain age gap (BAG) was calculated as the difference between corrected BA and chronological age: BAG = Corrected BA - Age. Our model predicted lower corrected BAGs in healthy controls compared to pwMS in the utilized cross-sectional sample (t_(1462)_=-19.40, p<2.2*10^-16^, Cohen’s d=-1.00, [-1.11,-0.89]). Moreover, we validated our models in BBSC data, showing within-subject correlations of Pearson’s r_sub1_=0.25, r_sub2_=0.23, r_sub3_=0.53 between age and brain age, yet with only the last correlation being significant (p_sub3_=0.006). Note that intra-class correlation coefficients could not be computed for these N=3 subjects due to the n-to-p ratio (small sample size, large number of observations). Additional information on the model validation procedure can be found at https://github.com/MaxKorbmacher/OFAMS_Brain_Age.

### Statistical Analysis

#### Descriptives

We first examined the validity of the stratification using linear mixed effects regression models with random intercept at the level of the individual to assess interaction effects between age and risk grouping, while controlling for age and sex. Specifically, age-group interaction effects on EDSS were used to evaluate the disability progression stratification, and age-group interaction effects on PASAT for processing speed decline stratification (Fig.1).

**Figure 1.** Sample stratification into functional and processing speed decline groups. Marginal interaction effects between group and age in random intercept models at the level of the individual indicate that risk groups (blue) show a faster functional and cognitive loss than functionally and cognitively stable or improving groups (red). Note that the stratifications were done independent from each other, resulting in n_PDG_=19 and n_PSDG_=11, with only n=3 participants being allocated in both groups.

To describe apparent differences at baseline, we analyzed univariate baseline differences between the respective risk vs stable groups using simple linear regression models, correcting for age and sex (Tables 1-2); and for differences in frequencies using Fisher’s exact tests (Supplemental Tables 5-6).

**Table 1:**
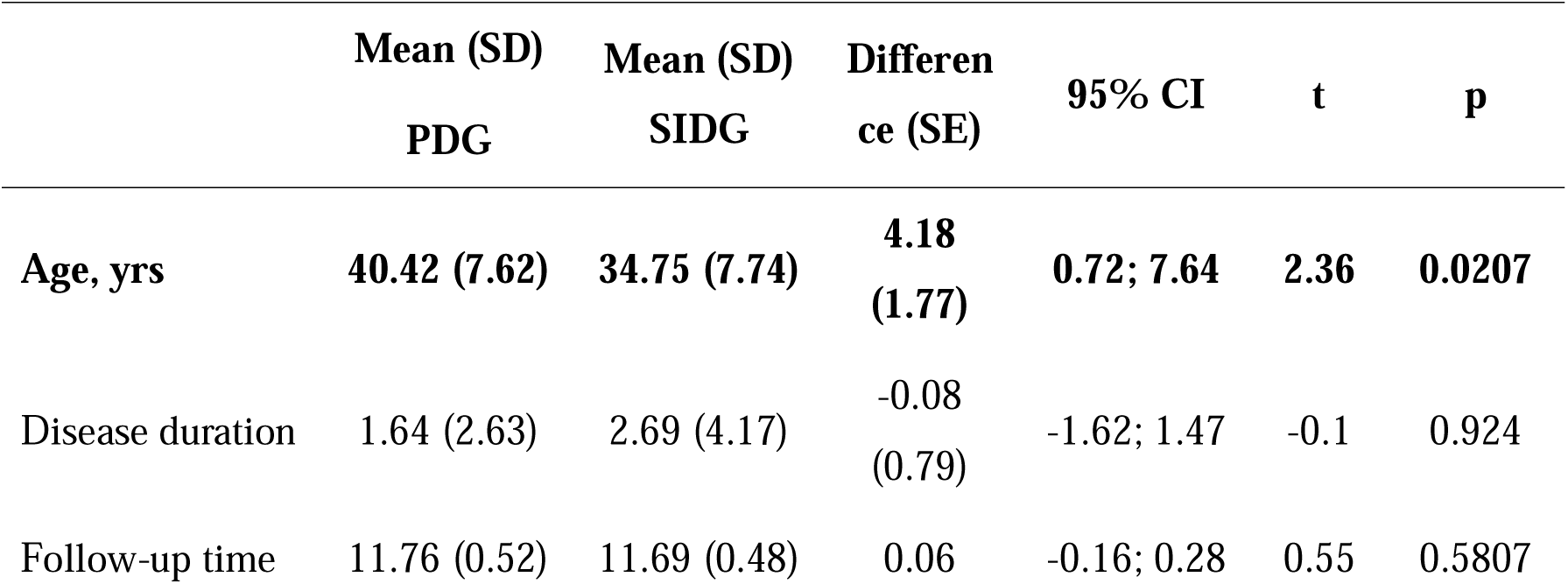
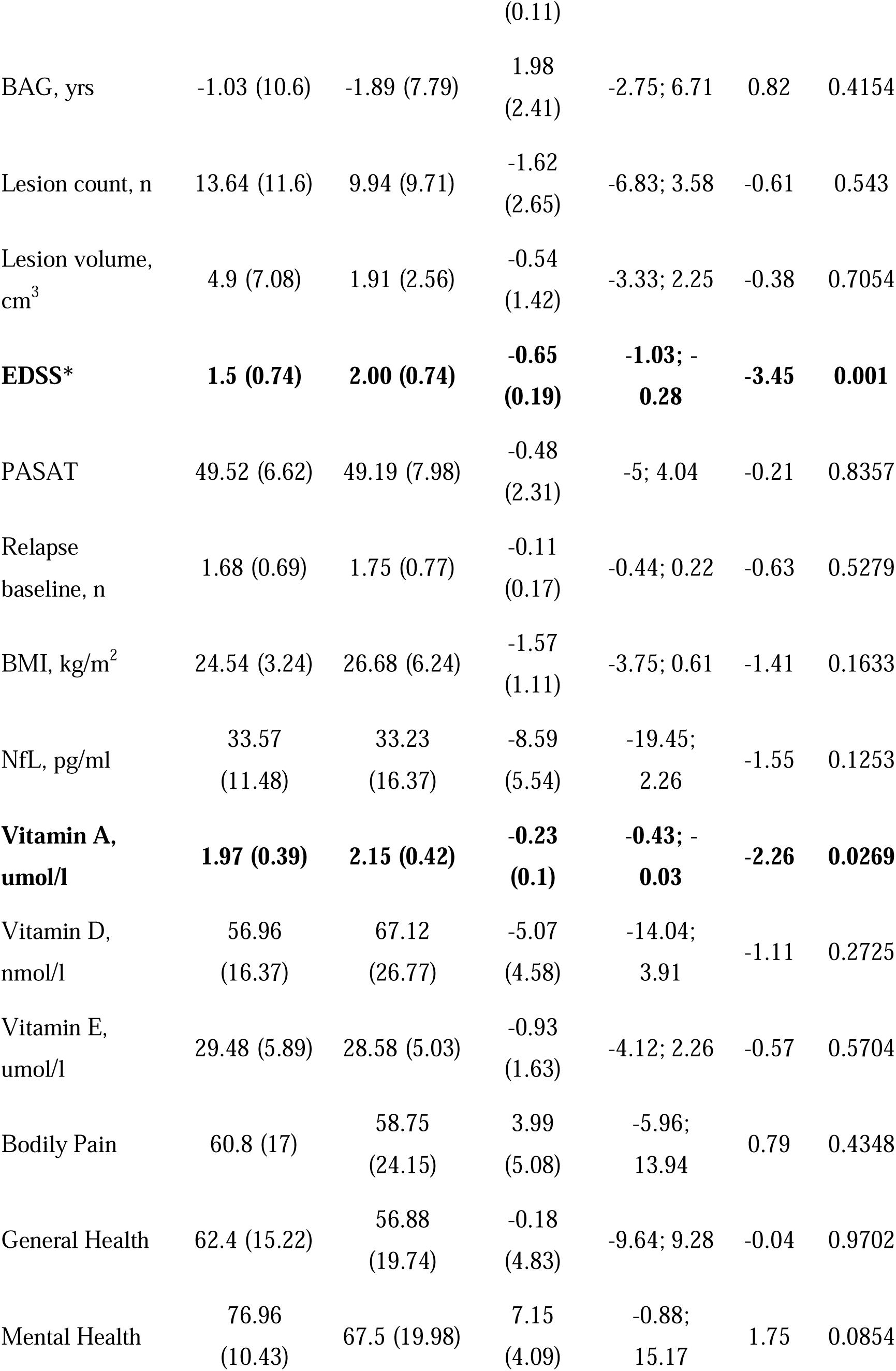
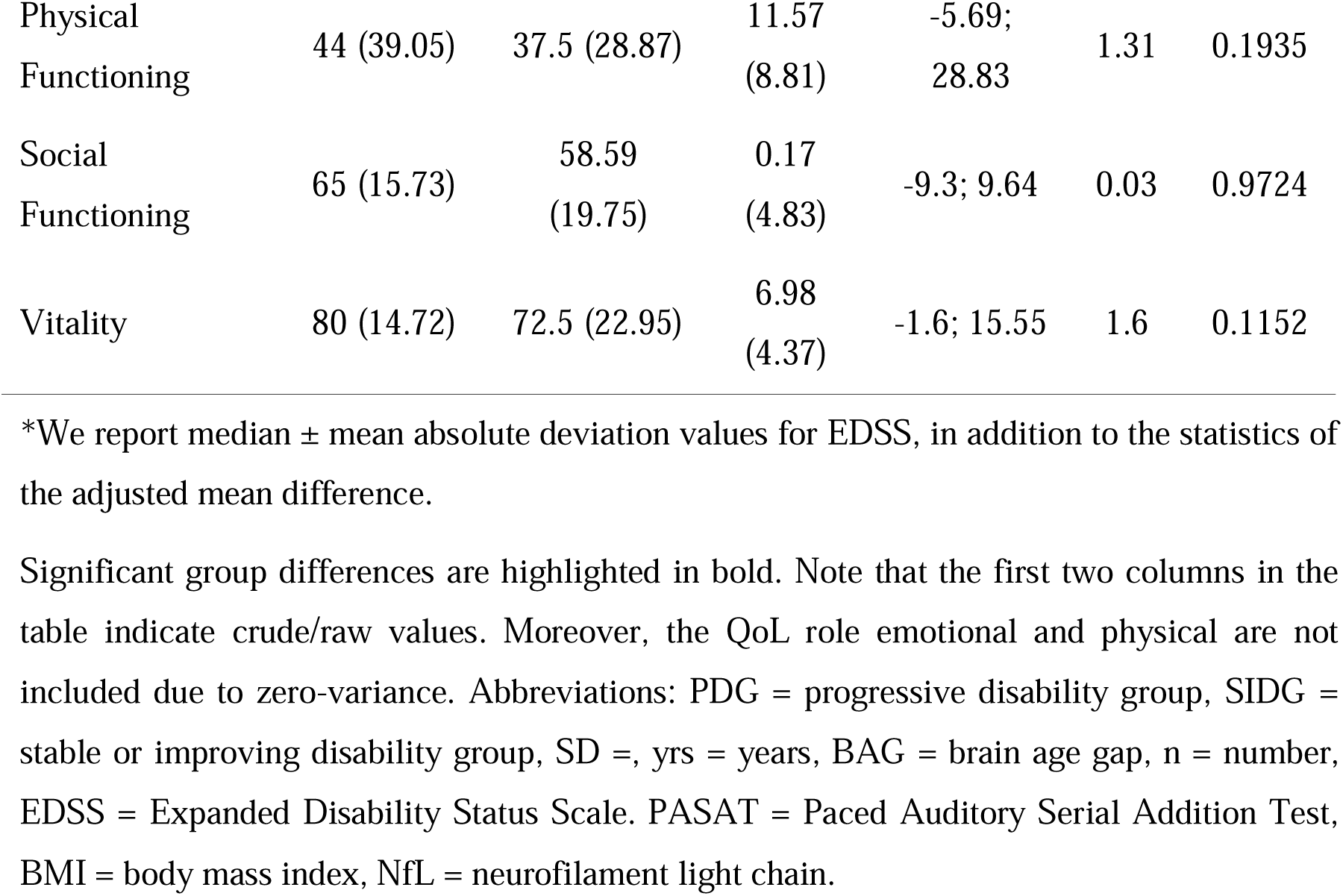
Baseline differences between disability progression and stable or improving disability groups. Continuous models were adjusted for age and sex, except for predictions of age, where only sex was used as covariate.

**Table 2:**
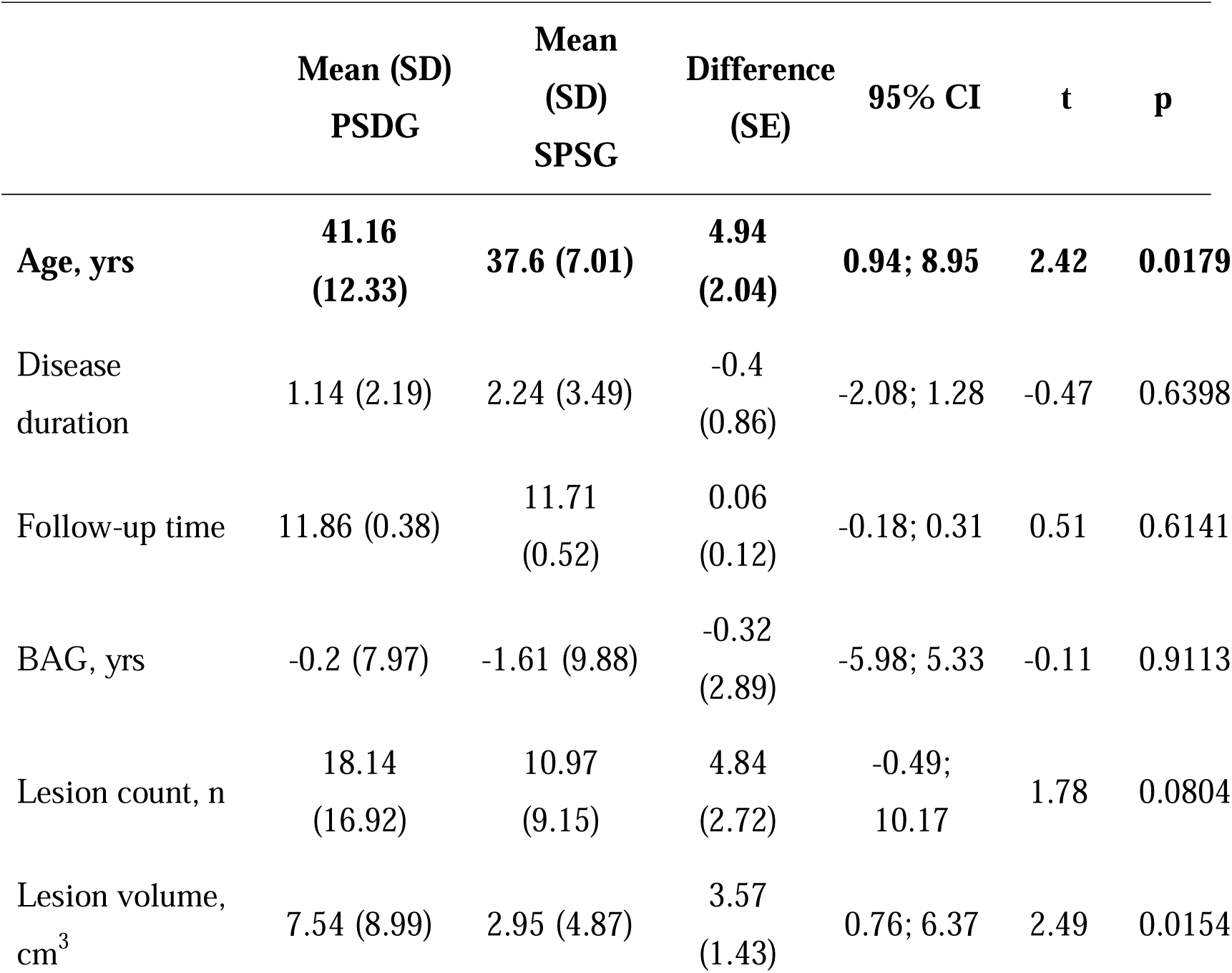
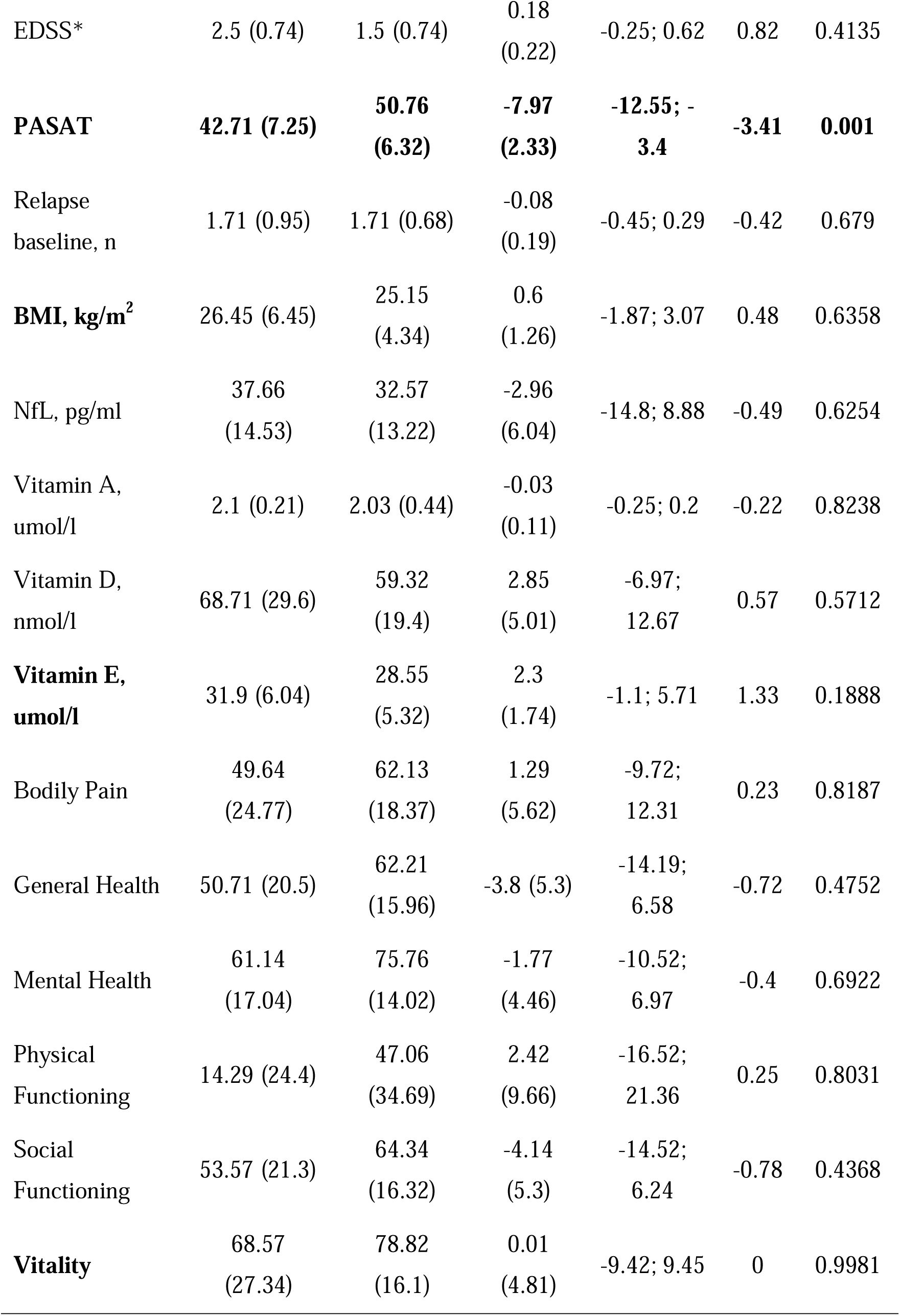
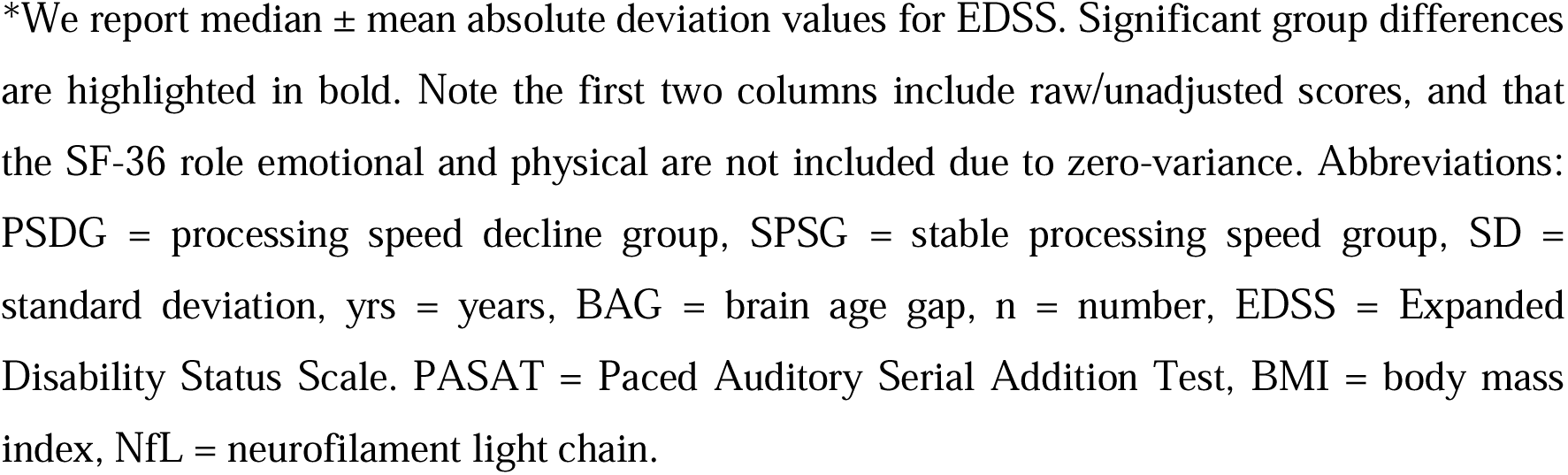
Baseline differences between processing speed decline and cognitively stable or improving groups. Continuous models were adjusted for age and sex, except for predictions of age, where only sex was used as covariate.

|  | Mean (SD)<br>PSDG | Mean<br>(SD)<br>SPSG | Difference<br>(SE) | 95% CI | t | p |
| --- | --- | --- | --- | --- | --- | --- |
| Age, yrs | 41.16<br>(12.33) | 37.6 (7.01) | 4.94<br>(2.04) | 0.94; 8.95 | 2.42 | 0.0179 |
| Disease<br>duration | 1.14 (2.19) | 2.24 (3.49) | -0.4<br>(0.86) | -2.08; 1.28 | -0.47 | 0.6398 |
| Follow-up time | 11.86 (0.38) | 11.71<br>(0.52) | 0.06<br>(0.12) | -0.18; 0.31 | 0.51 | 0.6141 |
| BAG, yrs | -0.2 (7.97) | -1.61 (9.88) | -0.32<br>(2.89) | -5.98; 5.33 | -0.11 | 0.9113 |
| Lesion count, n | 18.14<br>(16.92) | 10.97<br>(9.15) | 4.84<br>(2.72) | -0.49;<br>10.17 | 1.78 | 0.0804 |
| Lesion volume,<br>cm<sup>3</sup> | 7.54 (8.99) | 2.95 (4.87) | 3.57<br>(1.43) | 0.76; 6.37 | 2.49 | 0.0154 |
| EDSS* | 2.5 (0.74) | 1.5 (0.74) | 0.18<br>(0.22) | -0.25; 0.62 | 0.82 | 0.4135 |
| **PASAT** | **42.71 (7.25)** | **50.76**<br>**(6.32)** | **-7.97**<br>**(2.33)** | **-12.55; -**<br>**3.4** | **-3.41** | **0.001** |
| Relapse<br>baseline, n | 1.71 (0.95) | 1.71 (0.68) | -0.08<br>(0.19) | -0.45; 0.29 | -0.42 | 0.679 |
| **BMI, kg/m<sup>2</sup>** | 26.45 (6.45) | 25.15<br>(4.34) | 0.6<br>(1.26) | -1.87; 3.07 | 0.48 | 0.6358 |
| NfL, pg/ml | 37.66<br>(14.53) | 32.57<br>(13.22) | -2.96<br>(6.04) | -14.8; 8.88 | -0.49 | 0.6254 |
| Vitamin A,<br>umol/l | 2.1 (0.21) | 2.03 (0.44) | -0.03<br>(0.11) | -0.25; 0.2 | -0.22 | 0.8238 |
| Vitamin D,<br>nmol/l | 68.71 (29.6) | 59.32<br>(19.4) | 2.85<br>(5.01) | -6.97;<br>12.67 | 0.57 | 0.5712 |
| **Vitamin E,**<br>**umol/l** | 31.9 (6.04) | 28.55<br>(5.32) | 2.3<br>(1.74) | -1.1; 5.71 | 1.33 | 0.1888 |
| Bodily Pain | 49.64<br>(24.77) | 62.13<br>(18.37) | 1.29<br>(5.62) | -9.72;<br>12.31 | 0.23 | 0.8187 |
| General Health | 50.71 (20.5) | 62.21<br>(15.96) | -3.8 (5.3) | -14.19;<br>6.58 | -0.72 | 0.4752 |
| Mental Health | 61.14<br>(17.04) | 75.76<br>(14.02) | -1.77<br>(4.46) | -10.52;<br>6.97 | -0.4 | 0.6922 |
| Physical<br>Functioning | 14.29 (24.4) | 47.06<br>(34.69) | 2.42<br>(9.66) | -16.52;<br>21.36 | 0.25 | 0.8031 |
| Social<br>Functioning | 53.57 (21.3) | 64.34<br>(16.32) | -4.14<br>(5.3) | -14.52;<br>6.24 | -0.78 | 0.4368 |
| **Vitality** | 68.57<br>(27.34) | 78.82<br>(16.1) | 0.01<br>(4.81) | -9.42; 9.45 | 0 | 0.9981 |

\*We report median ± mean absolute deviation values for EDSS. Significant group differences are highlighted in bold. Note the first two columns include raw/unadjusted scores, and that the SF-36 role emotional and physical are not included due to zero-variance. Abbreviations: PSDG = processing speed decline group, SPSG = stable processing speed group, SD = standard deviation, yrs = years, BAG = brain age gap, n = number, EDSS = Expanded Disability Status Scale. PASAT = Paced Auditory Serial Addition Test, BMI = body mass index, NfL = neurofilament light chain.

#### Multiverse analyses

We used a multiverse approach to identify which baseline variables were most predictive of risk-membership using binary logistic regression (Figure 2). Multiverse analyses have the aim to test the robustness of effects using different analytical choices. Multiple models are run, leading to distributions of coefficients, instead of a single estimate. Hence, instead of presenting single coefficients, we present the central tendency of coefficents (median) and variability (median absolute deviation).

**Figure 2.** Multiverse estimates of predictors of disability progression. Note that disease modifying treatment at 12-year follow-up presented a strong effect of mOR=7.4±4.07 and was for readability not included.

Variables were excluded when the variance inflation factor (VIF) indicated an unacceptably high collinearity of VIF≥5 in a fully saturated model. These variables included SF-36 based general health and social functioning.

We included models with all possible combinations of variables of interest up to the 8^th^ order with age and a binary variable indicating whether disease modifying treatment was used at the 12-year follow-up as a covariates included in all models. All models were run with and without sex as a covariates, leading to a total of 213,522 models. The QoL “role limitations due to emotional problems” and “role limitations due to physical problems” were constant (mean=0±0) across participants and therefore excluded from the analyses.

McFadden’s^41^ pseudo R^2^ (pR^2^), the area under the curve (AUC) and Brier scores were estimated for each regression model as a model fit proxy. Wald-based power/sensitivity analysis^42^ was used to estimate the power of each model at α=0.05 by testing the assumption of a greater probability of disability progression or processing speed decline group-membership when predictors≠0. For the EDSS-based PDG, model fit was excellent (pR^2^=28.59%±8.00%; note that pseudo variance explained is generally lower than conventional variance explained^41^) when considering the 213,522 converged models, with 99.59% of the models having pR^2^>10%. The median power for these models was 20.82±30.89%. For the PASAT-based CDG the median model fit was excellent: pR^2^=40.00%±12.00% (100.00%>pR^2^>7.94%) considering the same converged models, presenting a medium power of nearly 0%. Considering the low median power, in 100 iterations producing each 10,000 datapoints, we simulated N = 1,000,000 datasets to allow for well-powered tests. For the procedure, we used classification and regression trees with the synthpop R package^43^. Note, that the data simulations entail the assumptions that the true population parameters can be found in the presented original data. These data were then used as a quality control set, running first a fully saturated model, then a model excluding SF-36 items, a model excluding MRI derivatives, and then a model only including significant variables or variables close to a median p-value=0.05 identified during the analysis of non-synthetic data, to assess whether potential redundancies of variable domains, comparing the models based on AIC, BIC, Brier score, area under the curve (AUC), and likelihood ratio tests.

Multiverse analyses ought to inform about all models, whether well specified or not. This is useful when assessing the literature while assuming analytic flexibility and the possibility of mis-specified models, both of which cannot be excluded. Hence, we reported on all models in the main text but reported only models selected for high power and/or excellent model fit in the supplement. We report raw/uncorrected p-values and 95% Confidence Intervals (CI) in square brackets when describing group differences for the stratifications. For multiverse analyses, median and median absolute deviation (MAD) are reported for the estimated ORs and p-values of each variable of interest across the specified models in the multiverse to a) belong to PDG vs SIDG or b) PSDG vs SPSG. Additionally, the proportion of the OR’s directionality (PORSD), such as the number of models including EDSS, with EDSS OR>1 for risk group membership, as share of all models was reported. When effects presented consistent directionality in 75% or more of the models they were used in, they were presented in the main text, in addition to clinically meaningful effect sizes. Clinically meaningful was defined here as plausible and possible changes based on the assessed scales and distributions of values and their changes, as well as the magnitude of the effect. For example, a 0.001% increase in the odds of belonging to one of the defined risk-groups per year of age at baseline might be a significant effect but accumulates to less than 1% higher odds even when being older than 100 years of age at baseline. Hence, in this example, the effect of age would not be counted as clinically meaningful. Finally, due to the relatively limited sample size and yet limited missingness, to avoid biasing the results, we did not conduct imputations of missing values. For baseline missingness of all utilized variables see Supplemental Figure 2, being relevant for the main analyses. EDSS and PASAT score missingness was relevant for stratification (see Supplemental tables 9-10).

## Results

### Study participants

The 85 pwMS with a 12-year follow-up were aged 38.9±8.3 (range: 19-58) years at baseline and 49.6±8.6 at the final follow-up, with 65.9% being females. Median baseline EDSS±MAD was 2.00±0.74, baseline PASAT was 47.9±9.4, and time since diagnosis was 1.9±3.2 years, and since the first symptom 5.5±5.5 years. Missingness was minimal (n≤6) for the baseline measures, which informed the main analyses (Supplemental Figure 2). Additional demographics and an overview of all variables can be found in the Supplemental Material. As an overview of the overlap between the different stratifications done in the following sections, note that the overlap between groups was small (n=9). 39 pwMS had favourable trajectories based on both stratifications, and 39 based on processing speed but not disability progression. 14 pwMS were identified as having a progressive disability but no cognitive decline, and only 12 pwMS had unfavourable trajectories based on both stratifications.

### Disability progression grouping

We show that, PDG presented a stronger increase in EDSS than SIDG (Figure 1), shown by a significant interaction effect between group membership and age on EDSS (p=7.12*10^-10^), indicating a 0.10±0.02 unit faster annual increase in EDSS in the PDG group.

At the same time, the groups were overall similar at baseline, with slightly better baseline disability, lower vitamin A levels, and higher age among the progressive disability group members (Table 1). There were no other significant differences (p>.05, Table 1), including differences in group sizes for sex, smoking, supplement/clinical trial treatment, risk genotype, or receiving disease modifying treatment at 12-year follow-up (Supplemental Table 5).

Significant group differences are highlighted in bold. Note that the first two columns in the table indicate crude/raw values. Moreover, the QoL role emotional and physical are not included due to zero-variance. Abbreviations: PDG = progressive disability group, SIDG = stable or improving disability group, SD =, yrs = years, BAG = brain age gap, n = number, EDSS = Expanded Disability Status Scale. PASAT = Paced Auditory Serial Addition Test, BMI = body mass index, NfL = neurofilament light chain.

### Multiverse analysis of disability progression

Models predicting a favourable/stable disability level presented a median area under the curve (mAUC) of 0.83±0.04, a median Brier score=0.16±0.02, and model fit of median pR^2^= 0.29±0.08, overall indicating good model perfermonce. Median effects which were on average significant (median p<0.05) were of relatively large size as well as unaffected by modelling choice in terms of their directionality (Figure 2, Supplemental Table 7). Varibles significantly predicting stable disability included receiving disease modifying treatment at 12-year follow-up (mOR_PDG_=7.44±4.07, p_median_=0.013, PORSD=100%), lower baseline EDSS (mOR_PDG_=0.25±0.11, p_median_=0.013, PORSD=100%), and counter-intuitively higher age (mOR_PDG_=1.12±0.04, p_median_=0.020, PORSD=100%), and lower vitamin A (mOR_PDG_=0.10±0.05, p_median_=0.016, PORSD=99.7%) and D levels (mOR_PDG_=0.95±0.02, p_median_=0.025, PORSD=100%).

### Processing speed decline grouping

The PSDG presented a stronger decrease in PASAT than SPSG (Fig. 1), shown by a significant (p=3.59*10^-5^) interaction effect between group membership and age on PASAT, indicating a 0.58-unit faster annual decrease in PASAT score in PSDG compared to SPSG. Moreover, at baseline, PSDG had higher age and PASAT score (Table 2). There were no other significant differences in continuous variables (p>0.05, Table 2), including differences in group sizes for sex, smoking, treatment, and risk genotype (Supplemental Table 6). However, the number of pwMS who received disease modifying treatment at the 12-year follow-up was lower among the PSDG (p<0.001; Supplemental Table 6).

### Processing speed decline multiverse analysis

Models predicting future processing speed decline presented strong predictive accuracy mAUC=0.89±0.05, Brier score=0.10±0.03, and excellent fit: median pR^2^= 0.40±0.12, but only receiving disease modifying treatment at follow-up (mOR_PSDG_=0.10±0.08, p_median_=0.013, PORSD=100%) and higher baseline PASAT score (mOR_PSDG_=0.86±0.03, p_median_=0.005, PORSD=99.73%) were significant predictors of future processing speed decline (Figure 3, Supplemental Table 8).

**Figure 3.** **Multiverse estimates of predictors of processing speed decline.**

### Simulation-based results

As a robustness check, we included analyses of N = 1,000,000 synthetic datasets, comparing fully specified models with models missing blocks of variables of MRI derivatives and SF-36 indices, and ultimately only including significant varibles for disability progression classifications and near-significant values for processing speed decline. For predictions of disability progression, the fully saturated models performed best (Table 3). Removing variable QoL/SF-36 and MRI domains or all but significant variables decreased the model performance. Differently, when predicting processing speed decline, removing QoL variables did only marginally change model performance. Removing MRI variables and removing all variables except for the ultimate disease modifying treatment and baseline PASAT score presented the largest effect on model evaluation metrics.

**Table 3:** Model evaluation metrics from selected models using simulated data.

| AUC | Brier score | AIC | BIC | Model | Prediction |
| --- | --- | --- | --- | --- | --- |
| 0.78 | 0.19 | 459489.37 | 459740.75 | Saturated | Disability Progression |
| 0.75 | 0.2 | 504289.15 | 504497.66 | SF-36 removed | Disability Progression |
| 0.76 | 0.2 | 821197.5 | 821426.73 | MRI removed | Disability Progression |
| 0.6 | 0.24 | 1229247.48 | 1229306.08 | only significant | Disability Progression |
| 0.73 | 0.16 | 427222.61 | 427475.3 | Saturated | Processing Speed Decline |
| 0.72 | 0.16 | 459631.47 | 459841.06 | SF-36 removed | Processing Speed Decline |
| 0.67 | 0.18 | 787135.32 | 787365.54 | MRI removed | Processing Speed Decline |
| 0.53 | 0.19 | 1116414.7 | 1116450.1 | only significant | Processing Speed Decline |

Abbreviations: AUC = area under the curve, AIC = Akaike information criterion, BIC = Bayesian information criterion.

Assessing the predictors in the fully saturated models reflected the predictor structure presented in the multiverse analysis. Predictors which were on average close to the common alpha-level of 0.05, as identified in the original data (disability progression: age, EDSS, disease modifying treatment, social functioning, vitality, vitamin D levels; processing speed decline: disease modifying treatment, PASAT), were also significant (p<0.001) and of the same direction in the fully saturated models based on the simulated data, with the exception of disease modifying treatment at follow-up indicating to increase the odds of favourable processing speed development, which contrasts the multiverse analysis, but is in line with the disability progression findings. The odds for stable disability group membership were 19% higher when receiving disease modifying treatmet at the 12-year follow-up (OR=1.19, 95% CI[1.17; 1.21]), by being older with 8% for every year (OR=1.08, [1.08; 1.08]). These odds were decreased with 53% for every 1-unit EDSS increase (OR=0.47, [0.46; 0.47]), by 17% for every umol/l increase in vitamin A (OR=0.83, [0.81; 0.84]) and by 1% for every nmol/l increase in vitamin D serum concentrations (OR=0.99, [0.99; 0.99]). The odds for stability in processing speed were increased by only 4% when receiving disease modifying treatmet at the 12-year follow-up (OR=1.04, [1.02; 1.25]), and decreased by 0.7% for every 1-point increase in baseline PASAT scores (OR=1.04, [1.02; 1.25]).

## Discussion

This study provides a comprehensive analysis of baseline variables associated with the risk and outcome of MS to assess their combined predictive ability for stratification of progressive disability (EDSS) and processing speed decline (PASAT). Our results are presented in the light of multiple possible analytical pathways, highlighting the influence of the selection of variables on the predictive value of each variable of interest in thousands of specified analytical paths.

Our analyses suggest a positive impact of transitioning towards disease modifying therapy, being the result of new drug discovery, for favourable disability progression. Higher baseline EDSS levels were however predictive of decreased odds of favourable outcomes. Counterintuitively, a higher baseline age as well as higher vitamin A and D serum concentrations indicated decreased odds for favourable disability progression. Finally, transitioning towards disease modifying therapy decreased the odds of favourable processing speed development, as well as higher baseline PASAT scores.

Our analyses are based on the identification of favourable compared to unfavourable disease trajectories. Such trajectories can be evaluated in different ways, often consulting EDSS scores which focus on physical function. However, previous evidence showcases that pwMS with functional disability progression do not necessarily correspond with those with comparably accelerated processing speed decline.^44^ The small overlap between the identified risk groups in our study confirmed this assumption. Moreover, quality controls ensured that the presented stratifications were robust to temporal fluctuations such as new relapses, new lesions or lesion activity, and we found clear differences in the trajectories when examining the variables which were stratified for.

Multiple studies have examined single or a small selection of MS markers.^17–21,25,27^ Yet only few studies have focussed on multivariate assessments and particularly on longitudinal data.^45–48^ Multimodal approaches have the advantage of enhancing predictions, for example as previously shown by combining electronic health records, clinical notes and neuroimaging data to predict MS severity^49^, conversion from clinically isolated syndrome to MS,^50^ or, similar to this study, to predict EDSS development.^51^ However, just as conventional statistical approaches, machine and deep learning techniques are influenced by the input variable selection and hyperparameter tuning suffers from generally small samples in the MS field.

The utilized input variables in our multivariate approaches have previously, often univariately, been associated with different MS outcomes. For example, higher EDSS scores^25^, HLA-DRB1 carriership^19^, lower vitamin levels^19–21^, higher lesion count^11^, cigarette smoking^22^, a lower PASAT test score^26^, several PROMs^27^, and a higher number of baseline relapses^24^ have previously been associated with progressive disability in MS, similar to elevated NfL levels^16–18^. CHI3L1 serum measures on the other hand were unstable in our sample (ICC≈0, Supplemental Figure 1). Hence, a meaningful interpretation of findings of higher CHI3L1 levels at baseline predicting less disability-progression at the 12-year follow-up not involving large uncertainty was not possible and we excluded the variable from our analyses. Cerebrospinal fluid-derived CHI3L1^17,18^ might be a more reliable measure. Additionally, our sample was relatively small and difficult to compare to more recent studies as patients were not treated during the first 6 months of the clinical trial. Hence, before drawing further inference, the influence of the characteristics of the examined limited sample and individual differences in disease trajectories need to be ruled out first by replicating of the findings reported here. Moreover, the statistical significance of the mentioned effects might depend on the selection of covariates. For example, the negative effect of cigarette smoking on health outcomes is generally known and has previously been documented in MS.^22^ However, to contribute to an acceleration of the disease or, for instance, functional or processing speed decline , other cumulative effects might need to be considered.

The efficacy of disease modifying treatment has been previously evidenced, with more recent therapies providing more efficient disease management via reduced relapse rate and disease progression.^52^ However, effects on cognitive impairment are rather unknown.^53^ Our findings are in line with these meta-analytic findings, where disease modifying treatments reduce increase the odds for a favourable disability progression. Their effects on cognitive impairment, measured here by processing speed, remain somewhat unclear, with our findings rather suggesting a negative effect of disease modifying treatments. While potential negative cognitive effects of switching medications as a response to new pharmacological developments, side effects, lack of effectiveness or necessary adjustments to the treatment plan cannot be excluded, the observed positive effects for the disability progression somewhat oppose this finding. Additional, due to the mentioned changes in treatment options and resulting policies, there is large variablility in treatment plans and administered drugs across the 12-year study.

We found a contraindicative effect of higher vitamin levels decreasing the odds of a positive disability trajectory. While this might be the case, there are multiple confounders relevant to this effect vitamin levels, such as immune function and nutritional health. Moreover, the examined vitamin A and D levels were within the recommended/reference range and far above what can be classified as deficiency.^54,55^

At the same time, as vitamins A and D present a broad influence on the immune system, baseline vitamin levels might indirectly reflect immune-related processes, where for example a lower serum level might indicate a larger metabolic need and hence an activated immune system.^56,57^ While vitamin D deficiency has been linked with a higher risk of developing MS,^19,36^ for example, vitamin D supplementation does not seem to significantly affect clinical outcomes in pwMS.^58^ Such associations are not established for other vitamin serum concentrations. Overall, the observed effects need further validation and replication in independent samples before being able to reach conclusion about their validity and meaning.

The main strengths of the study were the unique dataset, with near complete follow-up (95%) for over 12 years, in addition to a large brain age training set of more than 60,000 healthy subjects’ MRI data, and multiple validation sets, including hundreds of cross-sectionally scanned individuals. Combining these data allowed for more comprehensive analyses than previously conducted. Moreover, our analyses represent a good example for the potential of data-reusage. We were able to apply a new multiverse approach to the data, showcasing the influence of analysis pipelines on research outcomes.

Our study faces some limitations. First, several data-related issues limit the generalizability of the results. The examined historic data from a relatively homogenous Norwegian RRMS sample might bear little value for generalizations to other progressive MS or cohorts outside Northern Europe. At the same time, there was considerable variability of scanners and MRI protocols, which can influence the computation of metrics. This constitutes a major limitation. We decided against harmonisation procedures to avoid confounding effects, which are likely enhanced when the data size is limited.^59^

The size of the risk groups was relatively small, and treatment adjusted to the individual, including medication changes over time reduce the comparability of pwMS exacerbating the observation of true population estimates. Moreover, as common with automatic segmentation tools of MRI data, the segmentations were imperfect. MRI data processing challenges beyond the data processing software are scanner hardware and software changes over the acquisition period which could not be controlled for in the present national multi-centre study.

Machine learning implementations such as the utilized brain age model can “learn” and account for variability from unwanted covariates. Hence, the brain age predictions, which were trained on diverse, multi-site, software and field strength data, were meaningfully related to the examined developmental trajectories. The utilized brain age model is freely available together with a detailed user guide to enhance dissemination (https://github.com/MaxKorbmacher/OFAMS_Brain_Age).

Model misspecifications might have resulted from the thousands of probed modeling choices, including, for instance, not accounting for multicollinearity and non-linear relationships. On the other hand, the overall model performance was relatively strong and the directionality of significant effects consistent across analytic paths. Additionally, perfect model specifications were not the goal. Instead, we aimed to account for researcher bias, such as executing multiple but only reporting the significant analyses, while also testing multiple variables of interest in the same sample. This approach allows to closer approximate the reality of the literature than only models showing perfect fit.

We analysed a broad range of established and experimental MS markers. However, the selection of variables of interest in this study is not exhaustive and there might be more recently established markers which are more suitable for risk group stratifications, such as novel risk loci as DNM3–PIGC or DYSF–ZNF638^60^, giving additional inside into the heritability of MS.

The utilized brain age model focusses on cortical grey matter. While such approach improves the interpretability of the model, other approaches including subcortical grey matter^15^ or information on white matter microstructure^61^ might be more sensitive to MS-specific degeneration.

Overall, we show that pwMS can be stratified into disability progression and processing speed decline risk groups. The average predictive performance of the models was good. Predictors for such group stratifications, are dependent on analytic choices. Disability progression-relevant variables were the type of treatment at the 12-year follow-up (more recent disease modifying), EDSS, age, and potentially vitamin A and D levels, with further validations needed for these effects. Variables for the prediction of cognitive decline, measured by PASAT, were non-significant. Our findings offer not only additional insight into MS disease trajectories, but also a new perspective on analysing multiple MS markers. We encourage the MS research community to adopt the proposed multiverse approach to mitigate the influence of analysis pipelines on research outcomes and underline the potential for multivariate analyses. Additionally, we underscore the importance of incorporating disease stratifications, particularly using longitudinal data, as critical steps for advancing future studies.

## Data availability

Brain age model training data are available from the respective websites of the databases either openly or after application. OFAMS data can be shared after receiving a new ethics approval. Brain age models and analysis code are freely available at https://github.com/MaxKorbmacher/OFAMS_Brain_Age.

## Data Availability

Brain age model training data are available from the respective websites of the databases either openly or after application (see supplemental material). The main study data (OFAMS data) can be shared after receiving a new ethics approval upon reasonable request to the authors. Brain age models and analysis code are freely available at https://github.com/MaxKorbmacher/OFAMS_Brain_Age.

## Acknowledgements

We want to thank Nils Inge Landrø for the helpful comments and recommendations to improve this study.

We extend our sincere gratitude to the people with MS who participated in this study, whose contributions are invaluable. This research originated as a national intervention study (OFAMS) and ten years follow-up examinations. We thank the OFAMS study group at numerous hospitals across Norway.

Moreover, we want to extend our gratitude to the thousands of scanned participants in the numerous studies which served for establishing the brain age models, and the study facilitators.

## Contributors

Conceptualisation and design of the study: MK. Drafting the manuscript: MK. Statistical analyses: MK. Acquisition and analysis of data: All authors contributed in different capacities to data acquisition, administration, or analyses. Interpretation of data: MK. Critical revision of the article: all authors. Responsible for the overall content: MK. All authors approved the final version of the manuscript.

## Ethics Approvals

The study was approved by the Norwegian Regional Committees for Medical and Health Research Ethics (REK, 814351). The OFAMS-study and the 10-year follow-up were previously approved by REK (2016/1906) and registered as clinical trial (<u>clinicaltrials.gov</u> identifier: <u>NCT00360906</u>).

Ethical approval for the different brain age training datasets was obtained (REK 567301, PVO 17/21624), as well as for the longitudinal validation set (Bergen Breakfast Scanning Club, REK 238310), and the cross-sectional MS data (REK 2011/1846, REK 2016/102).

All participants gave their written informed consent. All methods were performed in accordance with the relevant guidelines and regulations (Declaration of Helsiniki).

## Supplemental Material

Supplemental material is available in a separate file.

