## Supplement for "Multimodal predictors of disability progression and processing speed decline in relapsing-remitting multiple sclerosis"

**Supplemental Material**

Supplemental material to the article “Multimodal predictors of disability progression and processing speed decline in relapsing-remitting multiple sclerosis” (Korbmacher et al., 2025).

**Supplemental Tables**

**Supplemental table 1: Overview of the collected variables**

| **Study month** | **0** | **1** | **2** | **3** | **4** | **5** | **6** | **7** | **8** | **9** | **12** | **18** | **24** | **144** |
| --- | --- | --- | --- | --- | --- | --- | --- | --- | --- | --- | --- | --- | --- | --- |
| **MRI** |  |  |  |  |  |  |  |  |  |  |  |  |  |  |
| T1Gd | X | X | X | X | X | X | X | X | X | X | X |  | X | X |
| T2 | X | X | X | X | X | X | X | X | X | X | X |  | X | X |
| T1-weighted 3D | |  |  |  |  |  |  |  |  |  |  |  |  | X |
| T2-3D sagital FLAIR | |  |  |  |  |  |  |  |  |  |  |  |  | X |
| **Clinical parameters** | |  |  |  |  |  |  |  |  |  |  |  |  |  |
| EDSS | X |  |  |  |  |  | X |  |  |  | X | X | X | X |
| Relapse recording | X | X |  | X |  |  | X | X |  | X | X | X | X | X |
| BMI | X | X |  | X |  |  | X | X |  | X | X | X | X |  |
| Tobacco use |  |  |  |  |  |  |  |  |  |  |  |  |  | X |
| **Genetics** |  |  |  |  |  |  |  |  |  |  |  |  |  |  |
| HLA-DRB*15 | X |  |  |  |  |  |  |  |  |  |  |  |  |  |
| WT1 | X |  |  |  |  |  |  |  |  |  |  |  |  |  |
| **PROMS** |  |  |  |  |  |  |  |  |  |  |  |  |  |  |
| Quality of life (SF-36) | X |  |  |  |  |  | X |  |  |  | X | X | X | X |
| **Cognition** |  |  |  |  |  |  |  |  |  |  |  |  |  |  |
| PASAT | X |  |  |  |  |  |  |  |  |  |  |  | X | X |
| **Laboratory** |  |  |  |  |  |  |  |  |  |  |  |  |  |  |
| Cotinine | X |  |  |  |  |  | X |  |  |  | X | X | X |  |
| alfa-tocopherol (vitamin E) | X | X |  | X |  |  | X | X |  | X | X | X | X |  |
| Retinol (Vitamin A) | X | X |  | X |  |  | X | X |  | X | X | X | X |  |
| Vitamin D | X | X |  | X |  |  | X | X |  | X | X | X | X |  |
| Serum-Neurofilament | X |  |  | X |  |  | X |  |  |  | X |  | X |  |
| Chitinase-3 | X |  |  | X |  |  | X |  |  |  | X |  |  |  |

**Supplemental table 2: Overview of datasets used for brain age model training**

| **Data** | **Sample size** | **Mean age ± SD, range** |
| --- | --- | --- |
| AddNeuroMed | 262 | 74±6.04, 53-90.1 |
| Adolescent Brain and Cognitive Development (ABCD) Study | 14,410 | 10.5±1.09, 8.92-13.5 |
| Healthy Brain Network (HBN) | 438 | 10.5±3.54, 5.11-21.2 |
| Human Connectome Project (HCP) | 767 | 28.4±3.57, 22-35 |
| Rockland Sample | 641 | 35.2±22.2, 6-85 |
| Tematisk Området Psykose (TOP) | 807 | 32.5±10.8, 13-72 |
| UK Biobank | 41,061 | 64.9±7.75, 46-83.7 |

*Note*, these values consider only the un-augmented training sample (n=58,317)

**Supplemental table 3: Overview of the MRI scanners and acquisition protocols**

| **Protocol (N of patients)** | **1 (3)** | **2 (13)** | **3 (3)** | **4 (1)** | **5 (3)** | **6 (4)** | **7 (3)** | **8 (5)** |
| --- | --- | --- | --- | --- | --- | --- | --- | --- |
| Scanner | Siemens Aera | Siemens Prisma | Siemens Skyra | Siemens Avanto | Siemens Skyra | Siemens Avanto | Siemens Aera | Philips Achieva |
| Field strength | 1.5T | 3T | 3T | 1.5T | 3T | 1.5T | 1.5T | 1.5T |
| Sequences | T1; MPRAGE T2; FLAIR | T1; MPRAGE T2; FLAIR | T1; MPRAGE T2; FLAIR | T1; MPRAGE T2; FLAIR | T1; MPRAGE T2; FLAIR | T1; MPRAGE T2; FLAIR | T1; MPRAGE T2; FLAIR | T1; FFE T2; FLAIR |
| TR (ms) | 1940 5000 | 1800 5000 | 2300 5000 | 2060 5000 | 2300 5000 | 2200 6000 | 2200 5000 | 7.6 4800 |
| TE (ms) | 2.69 335 | 2.28 386 | 2.32 387 | 3.10 340 | 2.32 387 | 2.82 358 | 2.67 335 | 3.75 338 |
| TI (ms) | 976 1800 | 900 1800 | 900 1800 | 1100 1800 | 900 1800 | 900 2200 | 900 1800 | 1660 1650 |
| Flip angle (¬∞) | 8 120 | 8 120 | 8 120 | 15 120 | 8 120 | 8 120 | 8 120 | 8 90 |
| Voxel size (mm) | 1.00x0.98x0.98 1.00x1.00x1.00 | 1.00x1.00x1.00 | 1.00x1.00x1.00 | 1.00x1.00x1.00 | 0.9x0.94x0.94 0.9x0.45x0.45 | 1.00x0.49x0.49 1.00x0.51x0.51 | 0.90x0.94x0.94 0.90x0.45x0.45 | 1.00x0.98x0.98 1.00x1.00x1.00 |
| **Protocol (N of patients)** | | **9 (12)** | **10 (2)** | **11 (6)** | **12 (1)** | **13 (2)** | **14 (8)** | **15 (3)** |
| Scanner | | Philips Achieva | Siemens Prisma | Philips Ingenia | Toshiba MRT200SP3 | Philips Ingenia | Siemens Prisma | Philips Achieva |
| Field strength | | 3T | 3T | 1.5T | 1.5T | 3T | 3T | 1.5T |
| Sequences | | T1; FFE T2; FLAIR | T1; MPRAGE T2; FLAIR | T1; FFE T2; FLAIR | T1; FFE T2; FLAIR | T1; FFE T2; FLAIR | T1; MPRAGE T2; FLAIR | T1; FFE T2; FLAIR |
| TR (ms) | | 8.04 5000 | 1800 5000 | 25 4800 | 13.5 1160 | 11.11 4800 | 1800 5000 | 7.1 4800 |
| TE (ms) | | 3.68 386 | 2.28 385 | 9.21 367 | 5.50 105 | 6.29 296 | 2.26 387 | 2.2 307 |
| TI (ms) | | 900 1800 | 900 1800 | 1660 2300 | 1650 906 | 1800 | 1800 | 1660 |
| Flip angle (¬∞) | | 8 90 | 8 120 | 30 90 | 20 90 | 8 90 | 8 120 | 8 90 |
| Voxel size (mm) | | 1.00x0.98x0.98 1.00x0.98x0.98 | 1.00x0.46x0.46 0.50x0.73x0.73 | 1.00x0.50x0.50 1.00x0.50x0.50 | 1.00x0.94x0.94 0.50x0.74x0.74 | 0.39x0.65x0.39 0.47x4.0x0.47 | 0.90x0.67x0.67 0.56x0.98x0.98 | 1.00x1.000x1.00 1.00x1.000x1.00 1.00x1.00x1.00 0.76x1.00x1.00 |

**Supplemental table 4: Brain age model validation metrics**

| **Sample and BA** | **Pearson's r** | **R^2^** | **MAE** | **RMSE** |
| --- | --- | --- | --- | --- |
| Training u | 0.9568 | 0.9154 | 5.0935 | 6.5013 |
| Training c | 0.9634 | 0.9281 | 4.8933 | 6.2186 |
| Test u | 0.9622 | 0.9259 | 5.2827 | 6.8292 |
| Test c | 0.9680 | 0.9370 | 5.2034 | 6.6508 |

BA = Brain Age, u = uncorrected, c = corrected, R2=variance explained, MAE = mean absolute error, RMSE = root mean squared error

**Supplemental table 5: Disability progression (DPG) and stable disability group (SDG) membership differences at baseline.**

|  | **DPG**  **Group membership = yes** | **DPG**  **Group membership = no** | **SDG**  **Group membership = yes** | **SDG**  **Group membership = no** | **OR** | **p** |
| --- | --- | --- | --- | --- | --- | --- |
| Sex (female = yes, male = no) | 22 | 14 | 26 | 14 | 0.85 [0.3,2.39] | 0.8134 |
| Smoking (yes, no) | 22 | 14 | 24 | 15 | 0.86 [0.3,2.4] | 0.8147 |
| ω-3 received (yes, no) | 17 | 21 | 13 | 18 | 1.52 [0.53,4.44] | 0.4691 |
| HLA−DRB1 carrier (yes, no) | 22 | 11 | 26 | 13 | 0.72 [0.24,2.14] | 0.6186 |
| Current treatment is disease modifying | 30 | 14 | 27 | 7 | 2.2 [0.7,7.47] | 0.2008 |

**Supplemental table 6: Processing speed decline group (PSDG) and stable processing speed (SPSG) membership differences at baseline.**

|  | **PSDG Group membership = yes** | **PSDG Group membership = no** | **SPSG**  **Group membership = yes** | **SPSG**  **Group membership = no** | **OR** | **p** |
| --- | --- | --- | --- | --- | --- | --- |
| Sex (female = yes, male = no) | 14 | 21 | 37 | 7 | 1.13 [0.36,3.87] | 1 |
| Smoking (yes, no) | 17 | 25 | 31 | 5 | 2.71 [0.81,10.73] | 0.1196 |
| ω-3 received (yes, no) | 6 | 27 | 26 | 13 | 0.48 [0.13,1.62] | 0.2821 |
| HLA−DRB1 carrier (yes, no) | 15 | 18 | 36 | 6 | 1.25 [0.37,4.61] | 0.7873 |
| Current treatment is disease modifying (yes, no) | 11 | 11 | 48 | 11 | 0.23 [0.07,0.76] | 0.0099 |

**Supplemental table 7: Favourable disability progression multiverse analysis results**

| **Names** | **Median OR** | **MAD OR** | **Median p** | **Portion OR>1** |
| --- | --- | --- | --- | --- |
| **Age** | **1.1238** | **0.0383** | **0.0195** | **1** |
| Bodily pain | 0.9853 | 0.0232 | 0.4597 | 0.24 |
| Body Mass Index | 0.9341 | 0.0654 | 0.3704 | 0.1513 |
| Brain Age gap | 1.0274 | 0.0311 | 0.5011 | 0.8154 |
| **Disease modifying treatment** | **7.442** | **4.0655** | **0.0132** | **0.9997** |
| **Expanded Disability Status Scale** | **0.2505** | **0.1119** | **0.0135** | **0** |
| HLA-DRB1 carrier | 0.6889 | 0.2807 | 0.5649 | 0.1558 |
| Lesion count | 0.9838 | 0.0247 | 0.6318 | 0.237 |
| Lesion volume | 1.0113 | 0.068 | 0.558 | 0.5658 |
| Mental health | 1.0342 | 0.031 | 0.2231 | 0.8745 |
| Neurofilament light chain pg/ml | 0.9684 | 0.0107 | 0.1434 | 0.0092 |
| Omega 3 supplement received | 1.6772 | 0.7652 | 0.4275 | 0.825 |
| Paced Auditory Serial Addition Test | 0.9816 | 0.0307 | 0.5258 | 0.245 |
| Physical functioning | 1.004 | 0.0106 | 0.5031 | 0.6371 |
| Relapses at baseline | 1.0581 | 0.3066 | 0.6702 | 0.5739 |
| Sex (female) | 0.7165 | 0.4845 | 0.4918 | 0.3025 |
| Smoking status | 1.2221 | 0.4703 | 0.6642 | 0.705 |
| **Vitality** | 1.0302 | 0.025 | 0.2536 | 0.8681 |
| **Vitamin A umol/L** | **0.1042** | **0.0496** | **0.0158** | **0.0026** |
| **Vitamin D nmol/L** | **0.9505** | **0.0165** | **0.0254** | **0** |
| Vitamin E umol/L | 0.9957 | 0.0534 | 0.5169 | 0.4628 |

Note that the results presented here include all possible analysis choices, including mis-specified models.

**Supplemental table 8: Favourable processing speed development multiverse analysis results**

| **Names** | **Median OR** | **MAD OR** | **Median p** | **Portion OR>1** |
| --- | --- | --- | --- | --- |
| Age | 1.0494 | 0.0452 | 0.3531 | 0.8492 |
| Bodily pain | 1.0054 | 0.029 | 0.5069 | 0.5915 |
| Body Mass Index | 1.1033 | 0.0745 | 0.2531 | 0.9381 |
| Brain Age gap | 0.9898 | 0.0687 | 0.541 | 0.4446 |
| **Disease modifying treatment** | **0.0988** | **0.075** | **0.0131** | **0** |
| Expanded Disability Status Scale | 1.5044 | 0.8686 | 0.4013 | 0.7462 |
| HLA-DRB1 carrier | 2.3315 | 2.0489 | 0.3817 | 0.8483 |
| Lesion count | 1.0625 | 0.0522 | 0.2034 | 0.8899 |
| Lesion volume | 1.2026 | 0.0855 | 0.0587 | 0.99 |
| Mental health | 0.9456 | 0.0477 | 0.1552 | 0.035 |
| Neurofilament light chain pg/ml | 0.9873 | 0.0159 | 0.4598 | 0.1969 |
| Omega 3 supplement received | 0.2223 | 0.1329 | 0.1308 | 0.0376 |
| **Paced Auditory Serial Addition Test** | **0.8585** | **0.0329** | **0.0045** | **0.0027** |
| Physical functioning | 1.0109 | 0.0165 | 0.305 | 0.7281 |
| Relapses at baseline | 0.6349 | 0.2526 | 0.4027 | 0.1317 |
| Sex (female) | 0.845 | 0.6671 | 0.592 | 0.4169 |
| Smoking status | 7.0069 | 4.6281 | 0.0536 | 0.988 |
| Vitality | 0.9935 | 0.0376 | 0.428 | 0.4223 |
| Vitamin A umol/L | 0.9601 | 0.7457 | 0.5559 | 0.4805 |
| Vitamin D nmol/L | 1.0321 | 0.0277 | 0.1447 | 0.967 |
| Vitamin E umol/L | 1.0192 | 0.0346 | 0.6888 | 0.7184 |

**Supplemental table 9: Missingness in EDSS scores across the entire study period**

|  | **All participants** | | **DPG** | | **SDG** | |
| --- | --- | --- | --- | --- | --- | --- |
| **Time point** | **Missing (total)** | **Missing (%)** | **Missing (total)** | **Missing (%)** | **Missing (total)** | **Missing (%)** |
| 0 | 4 | 4.71% | 0 | 0 | 4 | 6.06% |
| 6 | 7 | 8.24% | 0 | 0 | 7 | 10.61% |
| 12 | 4 | 4.71% | 0 | 0 | 4 | 6.06% |
| 18 | 5 | 5.88% | 1 | 5.26% | 4 | 6.06% |
| 24 | 6 | 7.06% | 1 | 5.26% | 5 | 7.58% |
| 144 | 7 | 8.24% | 0 | 0 | 7 | 10.61% |

Total N = 85, DPG N = 19, SDG N = 66

**Supplemental table 10: Missingness in PASAT scores across the entire study period**

|  | **All participants** | | **PSDG** | | **CSIG** | |
| --- | --- | --- | --- | --- | --- | --- |
| **Time point** | **Missing (total)** | **Missing (%)** | **Missing (total)** | **Missing (%)** | **Missing (total)** | **Missing (%)** |
| 0 | 0 | 0.00% | 0 | 0.00% | 0 | 0.00% |
| 24 | 2 | 3.03% | 0 | 0.00% | 2 | 3.39% |
| 144 | 3 | 4.55% | 1 | 4.55% | 2 | 3.39% |

PASAT data were only available for n=81 of the total N=85, i.e., there were n=4 systematically missing datasets. Hence the calculations in the table are based on a total sample of n=81, with n=22 PSDG members and n=59 CSIG members.

**Supplemental Figures**

**Supplemental Figure 1: Overview of the measurements of the key variables**.

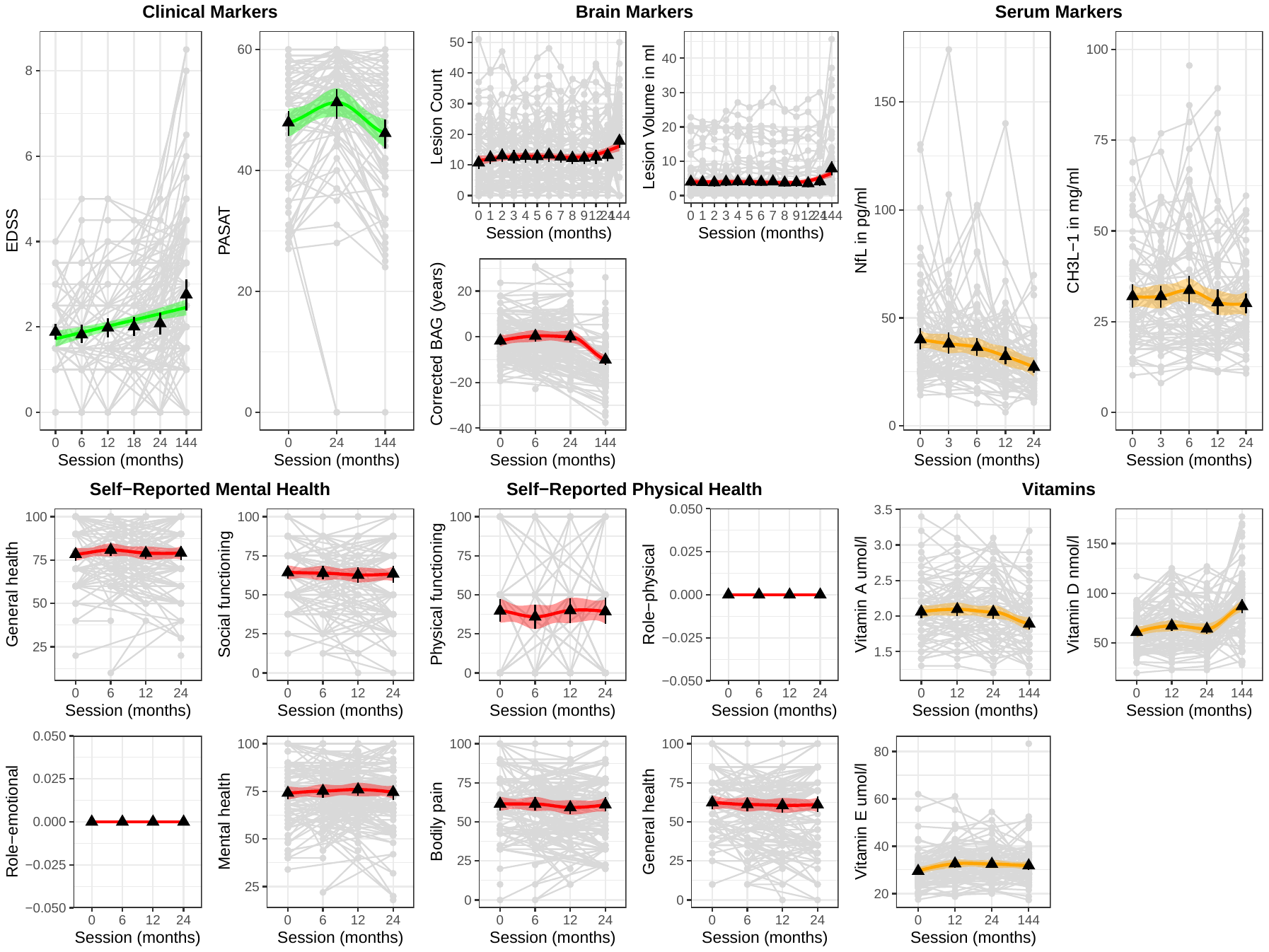

The grey dots represent data points per participant connected by grey lines. The triangles indicate mean values at each session with bootstrapped 95% confidence intervals. The lines indicate a smooth fit with 95% confidence interval for visual purposes. Note: a short-term training effect can be noted in PASAT scores.

**Intraclass correlation (ICC).** EDSS (ICC = 0.87 [0.83, 0.91], p = 3.9*10^-48^]) and PASAT (ICC = 0.86 [0.80, 0.90], p = 5.8*10^-26^]) measures showed strong associations across measurements. ICC for brain lesion count (ICC = 0.97 [0.96, 0.98], p = 7.2*10^-250^]) and volume (ICC = 0.99 [0.98, 0.99], p < 2.23*10^-308^]) were near-perfect, and good for the corrected brain age gap (ICC = 0.74 [0.63, 0.82], p = 1.6*10^-16^). While the ICC for NfL-levels was good, ICC = 0.83 [0.76, 0.88], p = 9.3*10^-35^, CH3L1-levels could not be reliably measured (ICC ≈ 0 [-0.38, 0.30], p = 0.49. The intra-class correlation coefficients were relatively high across measures. The quality-of-life measure with the lowest ICC=0.87 [0.82, 0.97], p = 4.1*10^-39^ was vitality. Among vitamins, vitamin D presented the lowest ICC = 0.56 [0.38, 0.69], p = 6.2*10^-7^, and vitamin A presented the largest ICC = 0.86 [0.80, 0.90], p = 1.4*10^-37^.

**Supplemental Figure 2: Missingness in the baseline (main analysis) variables**

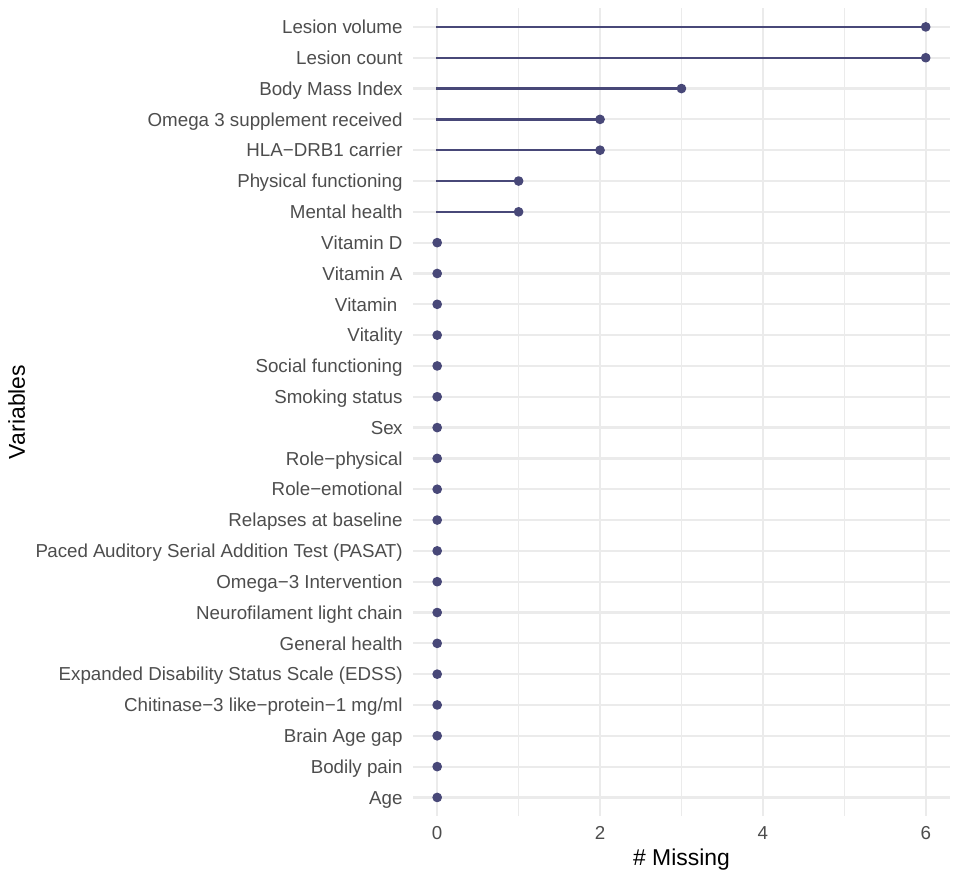
